# Evaluation of a camera-based monitoring solution against regulated medical devices to measure: Heart Rate, Respiratory Rate, Oxygen Saturation, and Blood Pressure

**DOI:** 10.1101/2022.10.20.22281306

**Authors:** Debjyoti Talukdar, Luis Felipe de Deus, Nikhil Sehgal

**Affiliations:** Department of Medical Research, Armenian Russian International University “Mkhitar Gosh”; Department of AI Research, Vastmindz Limited, London, United Kingdom

**Keywords:** heart rate, respiratory rate, oxygen saturation, blood pressure, camera based monitoring, medical device, remote photoplethysmography, rPPG

## Abstract

Regular monitoring of common physiological signs, including heart rate, blood pressure, and oxygen saturation, can be an effective way to either prevent or detect several potential conditions. In particular, cardiovascular diseases (CVDs) are a worldwide concern. According to the World Health Organization, 31% of all deaths worldwide are from CVDs. Recently, the COVID-19 pandemic has increased the interest in remote monitoring. At present, contact devices are required to extract most of an individual’ s physiological information, which can be inconvenient for users and may cause discomfort. However, remote photoplethysmography (rPPG) technology offers a solution for this issue, which enables contactless monitoring of the blood volume pulse signal using a regular camera, and ultimately can provide the same physiological information as a contact device. In this paper we propose an evaluation of rPPG technology against medical devices in a clinical environment, with a variety of subjects in a wide range of age, height, weight, and baseline vital signs. Results have shown that rPPG technology was able to meet the initial hypothesis of mean error of +/-3 units for Heart rate, Respiration Rate, and SpO2 estimation, as well as +/-10 mmHg for Systolic and Diastolic Blood Pressure.

## INTRODUCTION

There is growing interest in technologies related to remote patient monitoring (RPM) solutions, an interest that has largely piqued as of late amid the COVID-19 pandemic. However, not only COVID-19 has caused several deaths worldwide but also left uncountable consequences and side effects. Long before the pandemic, Cardiovascular Diseases (CVDs) have been desolating the world, according to the World Health Organization (WHO), 17.9 million people die each year from this condition, representing around 31% of deaths worldwide. CVDs are a group of health complications relating to the heart and blood vessels and include coronary heart disease, cerebrovascular disease, rheumatic heart disease, and other conditions [1].

It is therefore critical to regularly monitor some physiological parameters which can lead to early detection of such health complications.

Well-established methods for capturing physiological data include the use of the electrocardiogram (ECG) or photoplethysmography (PPG), both of which require the use of contact sensors. These methods, through their physiological signals, have the ability and the potential to measure several different physiological parameters such as heart rate (HR), respiration rate (RR), oxygenation (SpO2), and blood pressure (BP). Alternatively, researchers have recently introduced the remote-photoplethysmography (rPPG) technique which is a low-cost, non-contact method and an alternative solution for measuring the same parameters as the PPG signal in a contactless way. Since it is a method that can be performed on any consumer technology device with an embedded camera, its ease of use makes it an attractive addition to the suite of RPM solutions.

The information acquired through rPPG reflects the variations of blood volume in skin tissue which is modulated by cardiac activity. The reflection of light is influenced by the change in the volume of blood and of the movement of the wall of blood vessels, this phenomenon is visible within frame-to-frame changes of a red, green, and blue (RGB) camera. However, there are several challenges to extracting an optimal rPPG signal. Distortion in the signal might be caused due to low illumination, significant head movement, and the device properties in low-end devices such as the camera’ s frame rate and its resolution.

Any rPPG technology usually relies on a four-step methodology, which can be summarized as Frame-to-frame extraction, Region of Interest (ROI) detection, signal processing, and vitals estimation. Firstly, the video files are usually separated into several frames, the amount of frames in a certain period is denoted as frame rate, measured in frames per second (FPS).

There is, however, a minimum FPS required to pick up fast changes in the cardiac cycle. The regions of interest are theoretically compared to small sensors placed on the face and can diverge among authors, although a common step would be the detection of landmarks, which is performed by detecting face regions in each video frame. This process is commonly used with face-tracking algorithms such as the Viola-Jones method [2]. Once the ROIs are selected, pixel intensity components are extracted, and those components are in the RGB color space. In addition, the RGB components are spatially averaged over all pixels in the ROI to yield a red, blue, and green component for each frame and form the raw signals.

Furthermore, the signal processing stage is applied, also known as the “rPPG Core”. The rPPG Core has been the object of various studies in the last decade, resulting in multiple methods which seek to extract the signal as clean as possible from RGB components. Some rely on Blind Source Separation (BSS) methods, which can retrieve information by de-mixing raw signals into different sources. Principal Component Analysis (PCA)-based and Independent Component Analysis (ICA)-based, which use different criteria to separate temporal RGB traces into uncorrelated or independent signal sources are some of the techniques used [3-4]. Other authors have tried to improve the quality of the signal by changing the color space to a chrominance-based domain [5].

In a more recent study, Wang et al. [6] introduced a new alternative to process RGB components into rPPG signal, called the “plane-orthogonal-to-skin” (POS) algorithm. The algorithm’ s main idea is to filter out the intensity variations by projecting RGB components on a plane orthogonal to a determined normalized skin tone vector. As a result, a 2-D signal referent to the projections is obtained and then combined into a 1-D signal which is one of the input signal dimensions that is weighted by an alpha parameter. The alpha parameter is the quotient of the standard deviations of each signal.

## MATERIALS AND METHODS

This study aimed to compare the accuracy of remote health screening technology against gold standard vital signs monitors (hereon referred to as the reference method) in the measurement of HR, RR, BP, and Sp02. The study involves accuracy evaluation of heart rate, respiratory rate, oxygen saturation and blood pressure using Vastmindz’ s 3.0 SDK that can be integrated into android and iOS mobile apps as well as Web apps and provides an estimation of physiological assessments for a variety of vital signs. It is worth considering that the reference instruments used also have some degree of error and in some cases can deviate by up to +/-3 units per parameter. This will be taken into account when discussing the results.

### Methods

The methods of this study are divided into two blocks, the onsite data collection, and the analysis. The data was collected at the hospitals without any interference from any staff. The device used in data collection was running an app with Vastmindz’ s technology, the app interface will effectively perform face detection, place landmarks, and create ROIs, after that BGR components of each ROI within each frame will be collected and stored in a cloud database. The BGR samples are the only visual information stored, Fig.2 shows an overview of the method.

**Figure 1:**
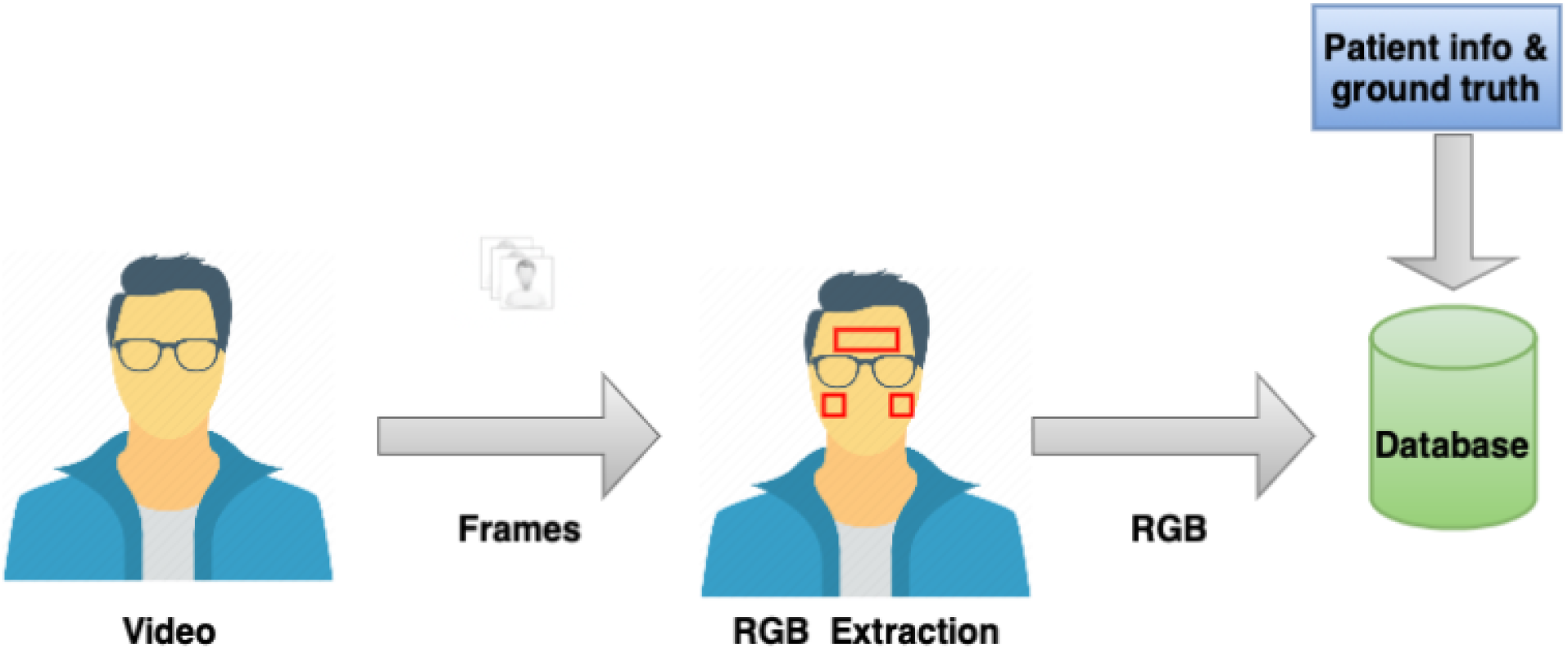
Study methodology - Data collection

**Figure 2:**
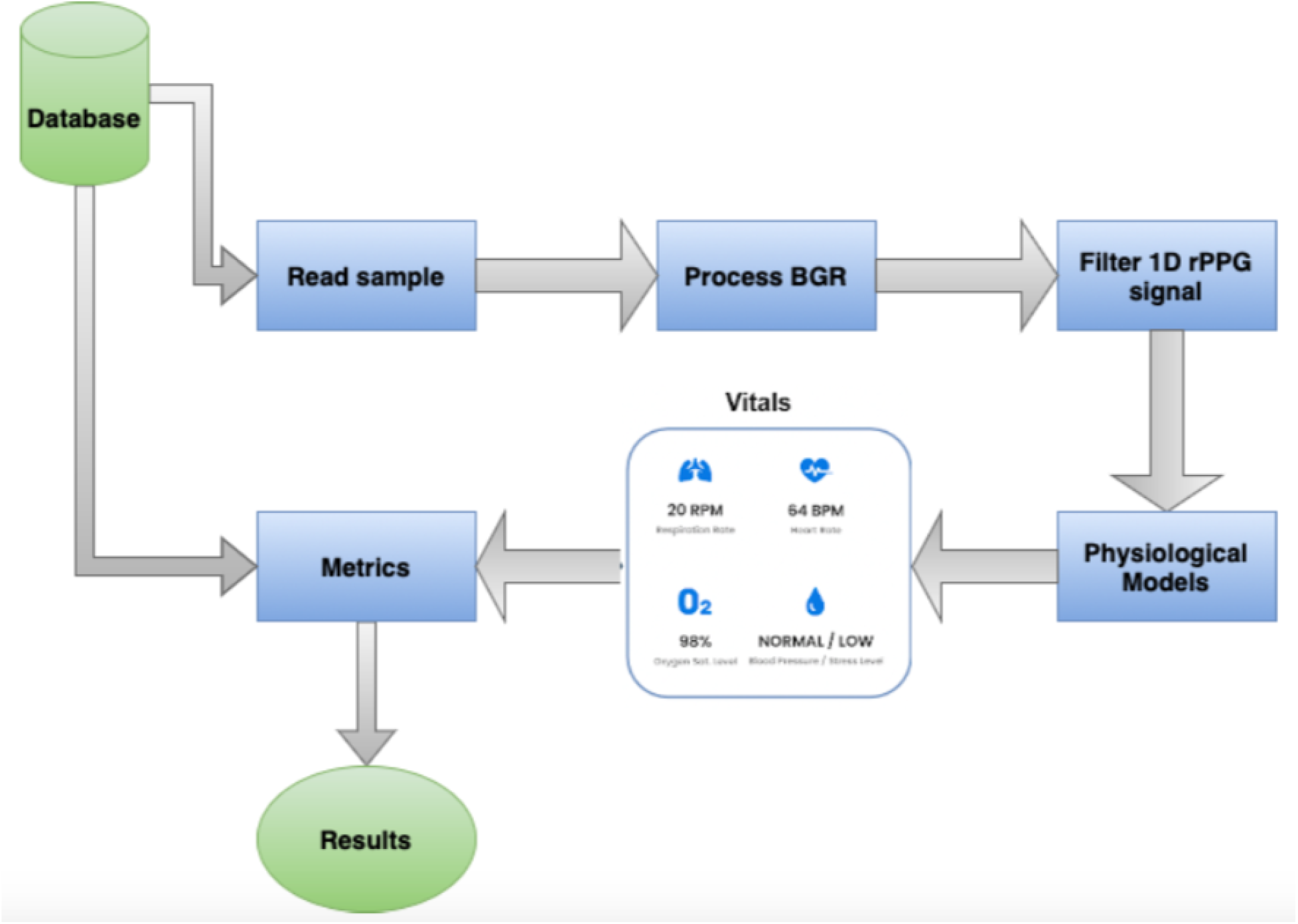
Study methodology - Analysis

Using the data collected, a pipeline was built to first, parse the sample, process the BGR components into a 1D signal, and extract each one of the physiological parameters. Lastly, the results extracted were then compared to the ground truth obtained with medical devices. Using the comparison between ground truth and estimated values, the metrics Mean Error (ME), Mean Absolute Error (MAE), Root Mean Squared Error (RMSE), and Root Mean Squared Percentage Error (RMSPE %) were calculated. Fig.3 summarizes the workflow [7].

**Figure 3:**
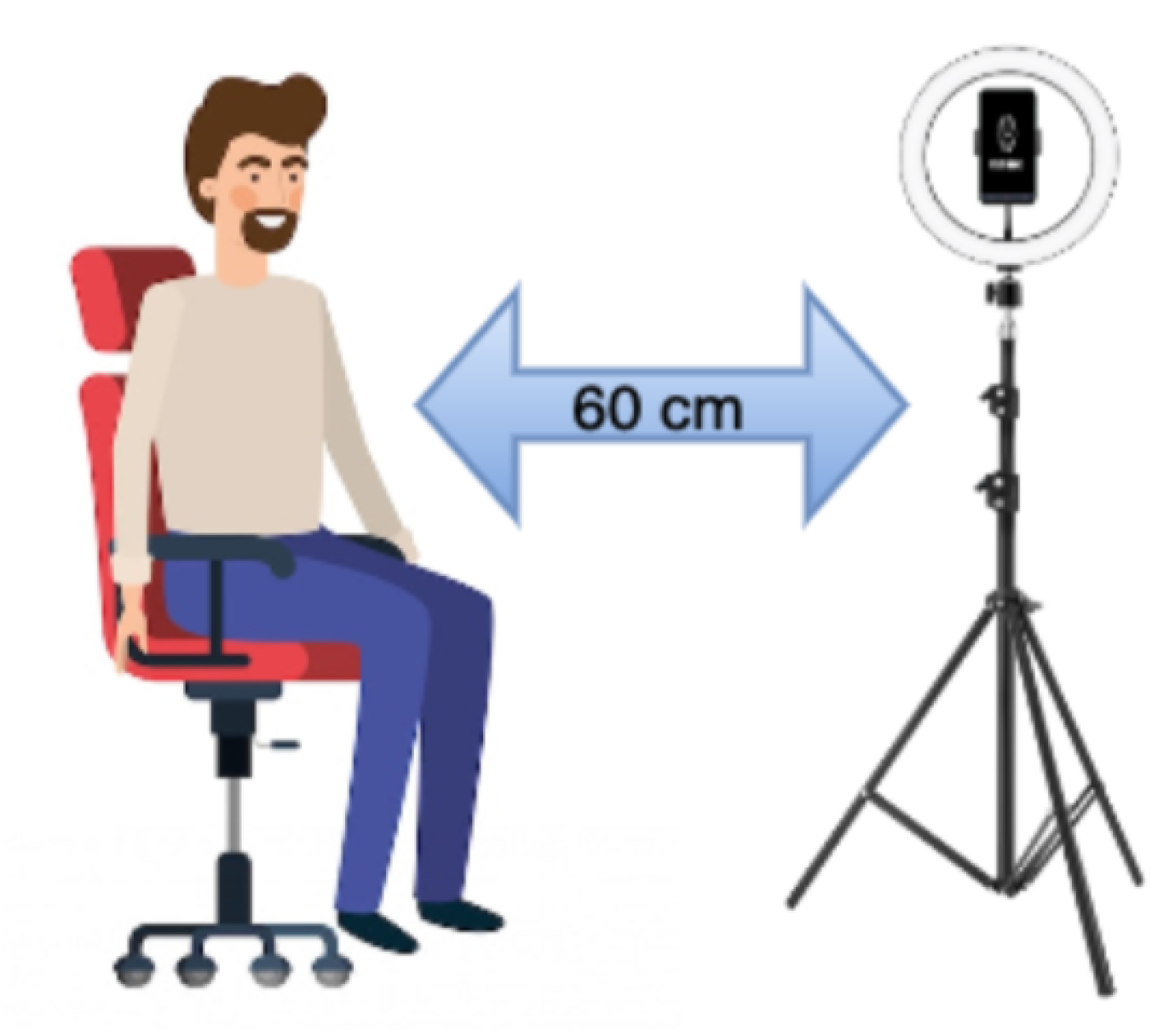
Study methodology - Protocol

### Study design

Five vital signs, Heart Rate, Respiration Rate, Oxygen Saturation, Systolic Blood Pressure (SBP), and Diastolic Blood Pressure (DB), were simultaneously measured using medical devices. In the study, multiple devices were used: an HP laptop with Logitech webcam, Smartphones: Samsung M32, One Plus 9, iPhone 11, and an iPad 9. The idea was to diversify the device to show that the results do not suffer from any technology bias, working in Android, iOS, Windows, and macOS systems [8].

The device was positioned 60 cm away from the subject’ s face and fixed to a tripod with a ring light source in front of the subject (minimum of 250 lux). Reference measurements were extracted with: Massimo mightiest for HR and RR; OMRON / CIRCA MICRO LIFE/ NIDEK Multi-parameter monitor and pulse oximeters. Subjects were asked to remove glasses or any other facial coverings and to be seated in a chair. During the scan, subjects were instructed to remain completely still and look straight at the camera during the entire duration of the scan.

Each scan took 60 seconds and all the vitals were measured once using the medical devices. Fig.1 shows the experiment setup [9].

## RESULTS

A total of 463 subjects were recruited for the study. 179 Females and 287 Males, aged between 18-83 (42), with various skin colors, ethnicities, and medical statuses participated in this study. A summary of patient demographic characteristics can be found in Table 1.

**Table 1:** Table of Demographic characteristics

| Characteristics | Mean (min - max) |
| --- | --- |
| Age | 42.07(18 - 83) |
| BMI | 25.11(7 - 34) |
| Height | 165.29(147 - 252) |
| Weight | 68.45(32 - 103) |

Aside from evaluating the technology in a variety of patients with different demographic information, such as age range, height, and weight, it is also important to analyze subjects with different baseline parameters, such as resting pulse rate, respiration rate and blood pressure. An overview of the reference values can be seen in Table 2.

**Table 2:** Table of Vital signs ground truth

| Vital sign | Mean (min - max) |
| --- | --- |
| Heart Rate (HR) | 80.36(51 - 124) |
| Oxygen Saturation (SpO2) | 97.57(91 - 100) |
| Respiration Rate (RR) | 17.62(8 - 36) |
| Systolic Blood Pressure (SBP) | 125.81(82 - 205) |
| Diastolic Blood Pressure (DBP) | 80.32(50 - 119) |

The reference values for each subject were compared to the estimated through facial scanning. Table 3 shows for each vital sign the sample size, mean error, mean absolute error, root mean squared error and root mean squared percentage error. Moreover, results for each vital sign were split into three groups, to present results within separated classes. Apart from the numeric values in the above table, it is also possible to evaluate the results through charts that provide a different view of the correlations. Fig.4 shows histograms of the errors for each one of the vital signs for a) Respiration Rate, b)Oxygen Saturation and c) Heart Rate. Whilst Fig.5 shows the same plot for Blood Pressure, in terms of a) Systolic and b) Diastolic.

**Table 3:**
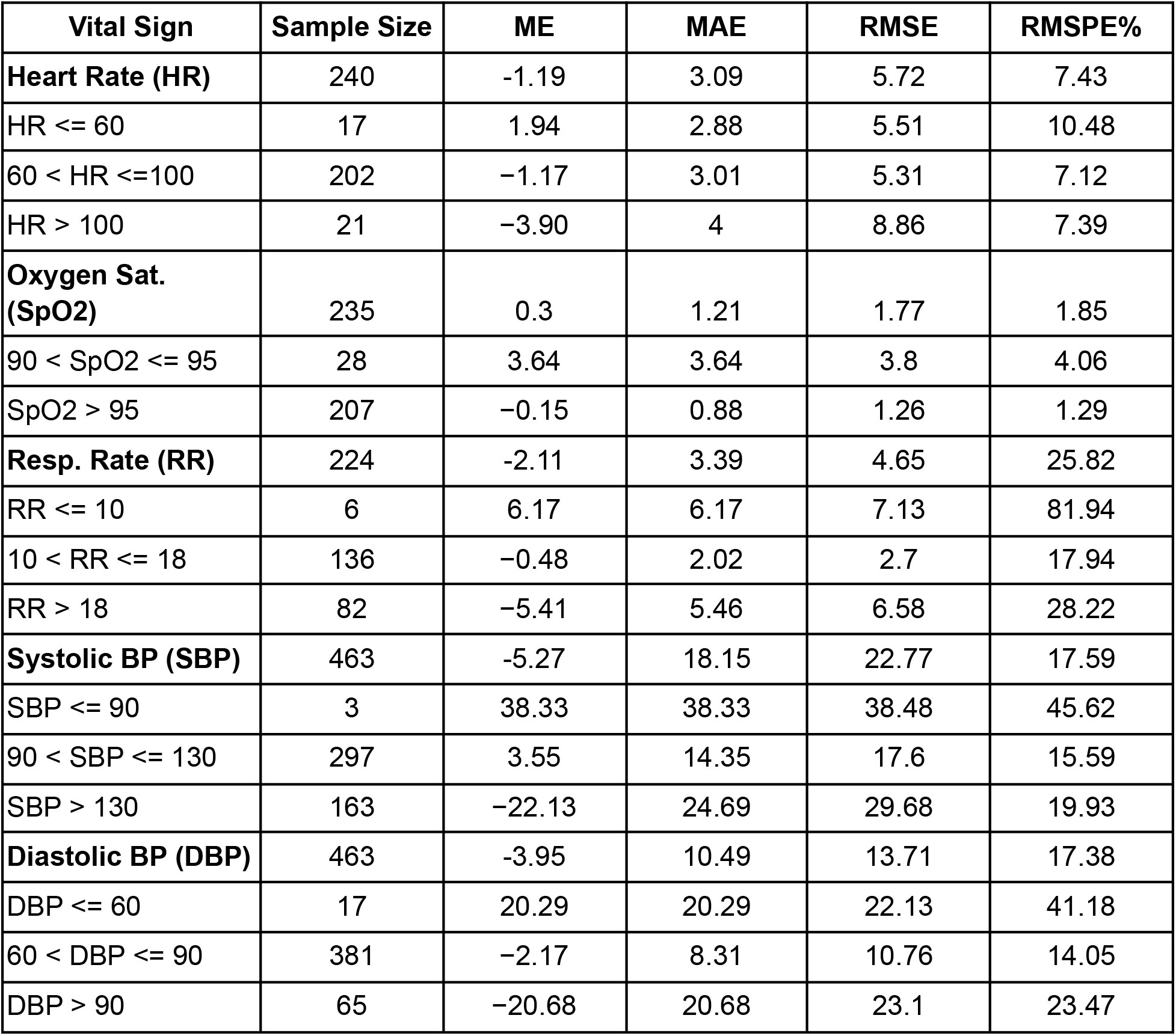
Table of differences between reference and estimated

| **Vital Sign** | **Sample Size** | **ME** | **MAE** | **RMSE** | **RMSPE%** |
| --- | --- | --- | --- | --- | --- |
| **Heart Rate (HR)** | 240 | -1.19 | 3.09 | 5.72 | 7.43 |
| HR <= 60 | 17 | 1.94 | 2.88 | 5.51 | 10.48 |
| 60 < HR <=100 | 202 | -1.17 | 3.01 | 5.31 | 7.12 |
| HR > 100 | 21 | -3.90 | 4 | 8.86 | 7.39 |
| **Oxygen Sat. (SpO2)** | 235 | 0.3 | 1.21 | 1.77 | 1.85 |
| 90 < SpO2 <= 95 | 28 | 3.64 | 3.64 | 3.8 | 4.06 |
| SpO2 > 95 | 207 | -0.15 | 0.88 | 1.26 | 1.29 |
| **Resp. Rate (RR)** | 224 | -2.11 | 3.39 | 4.65 | 25.82 |
| RR <= 10 | 6 | 6.17 | 6.17 | 7.13 | 81.94 |
| 10 < RR <= 18 | 136 | -0.48 | 2.02 | 2.7 | 17.94 |
| RR > 18 | 82 | -5.41 | 5.46 | 6.58 | 28.22 |
| **Systolic BP (SBP)** | 463 | -5.27 | 18.15 | 22.77 | 17.59 |
| SBP <= 90 | 3 | 38.33 | 38.33 | 38.48 | 45.62 |
| 90 < SBP <= 130 | 297 | 3.55 | 14.35 | 17.6 | 15.59 |
| SBP > 130 | 163 | -22.13 | 24.69 | 29.68 | 19.93 |
| **Diastolic BP (DBP)** | 463 | -3.95 | 10.49 | 13.71 | 17.38 |
| DBP <= 60 | 17 | 20.29 | 20.29 | 22.13 | 41.18 |
| 60 < DBP <= 90 | 381 | -2.17 | 8.31 | 10.76 | 14.05 |
| DBP > 90 | 65 | -20.68 | 20.68 | 23.1 | 23.47 |

**Figure 4:**
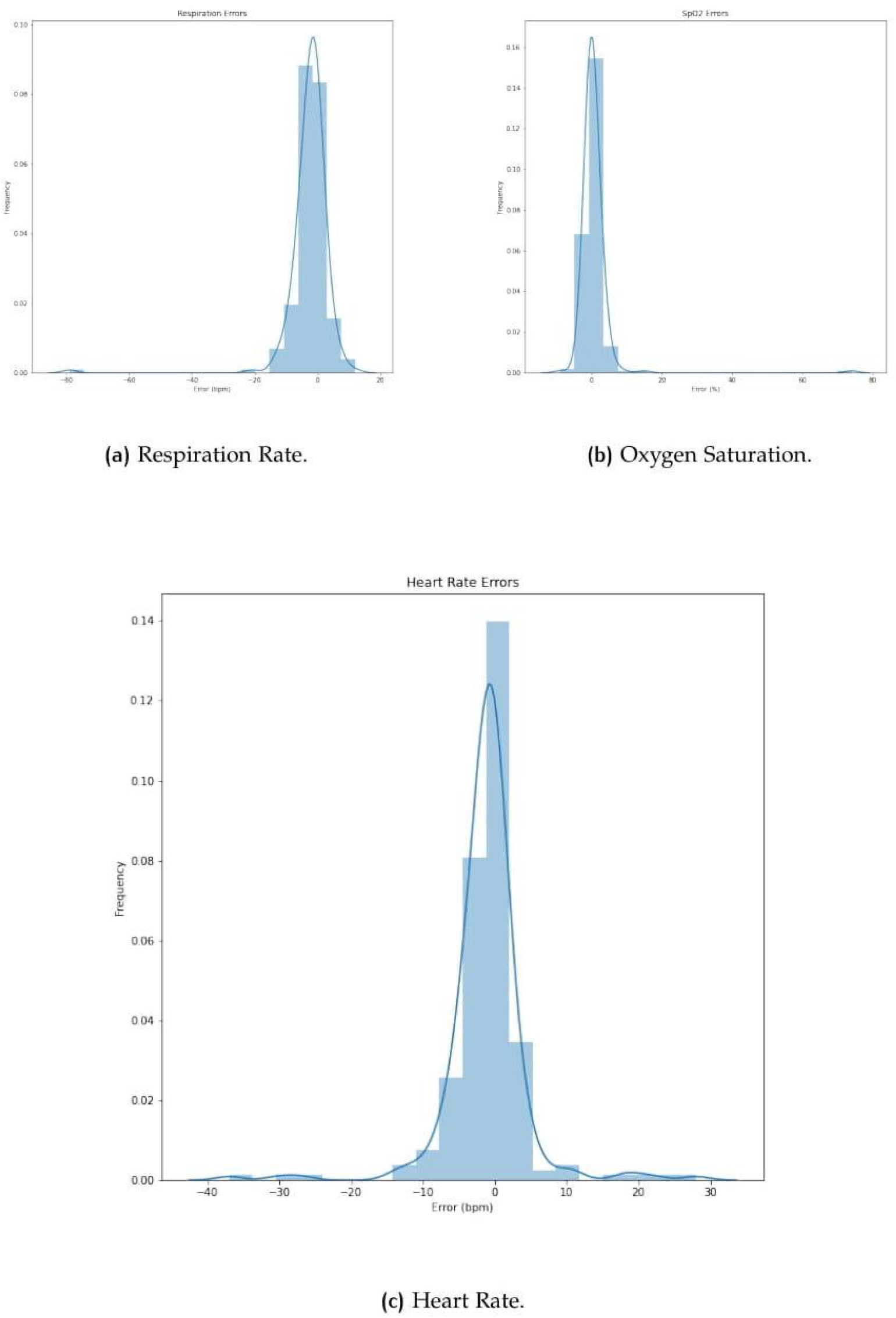
Histograms of errors for: a) Respiration Rate, b) Oxygen Saturation and c) Heart Rate

**Figure 5:**
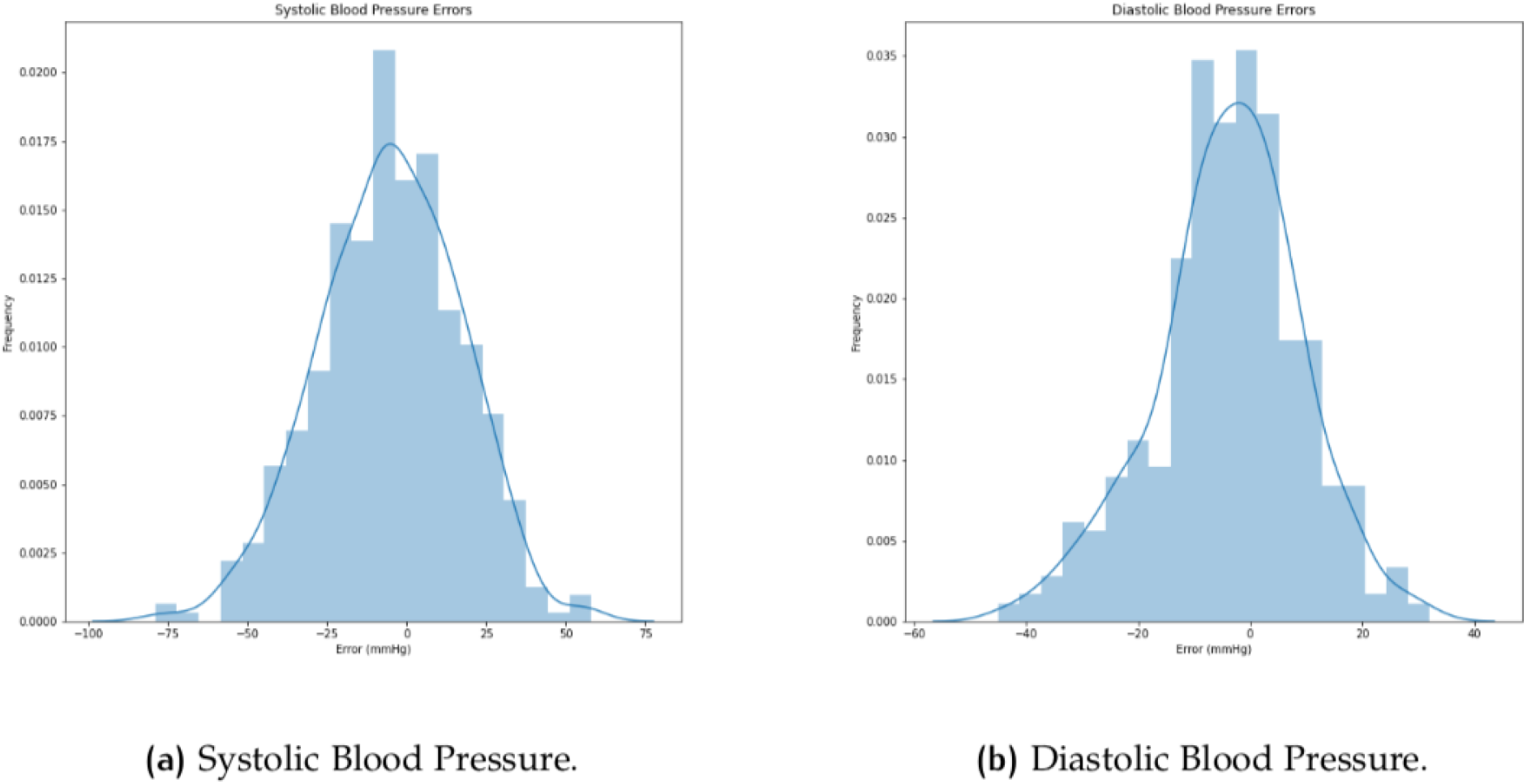
Histograms of errors for Blood Pressure

Despite histograms, which show the error distribution, it is also possible to evaluate through Bland Altman plots, which show the results in terms of mean value vs. difference for each data point. Fig.6 and Fig.7 show the plots for all the vitals.

**Figure 6:**
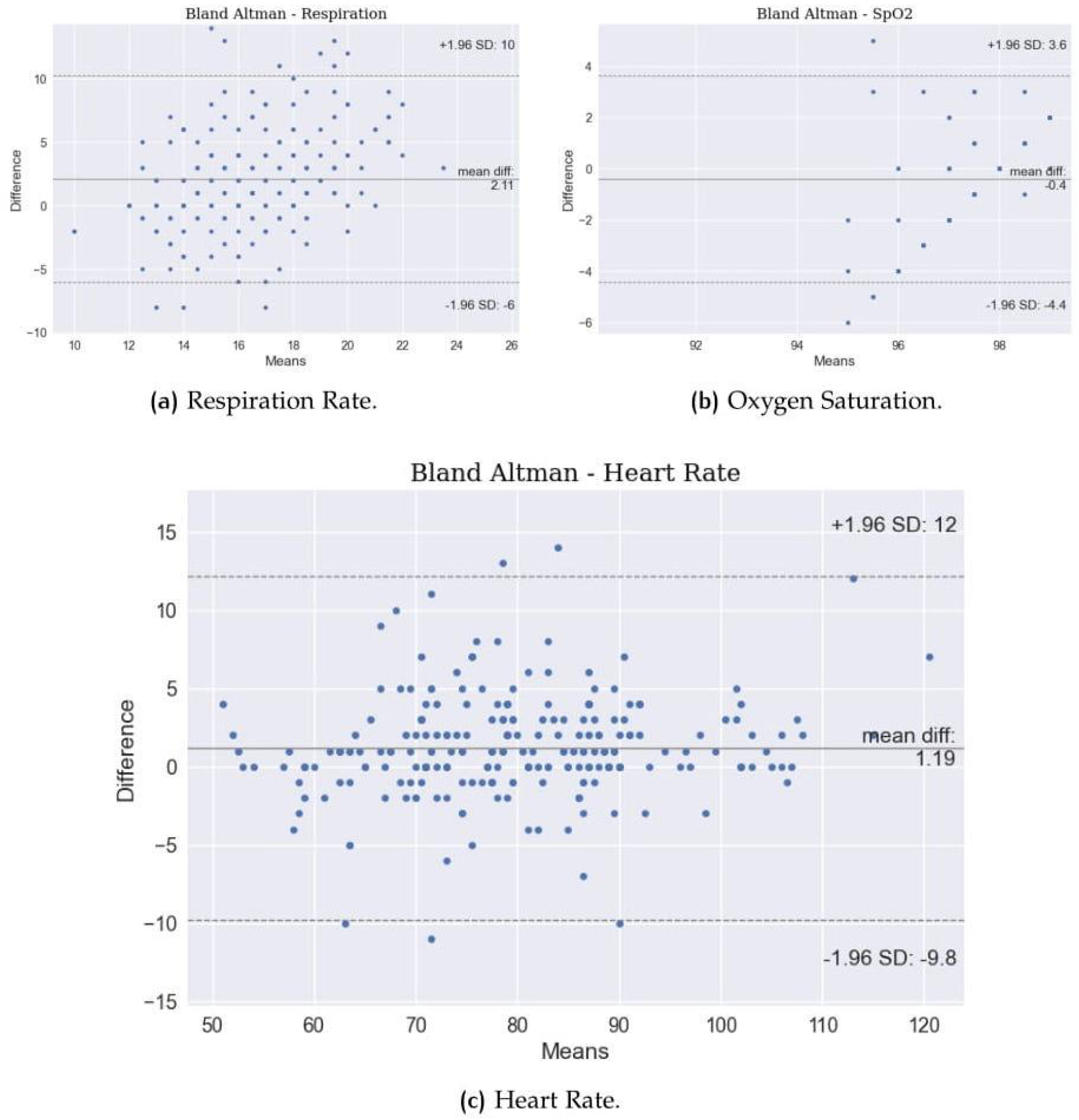
Bland Altman Plot for: a) Respiration Rate, b) Oxygen Saturation and c) Heart Rate

**Figure 7:**
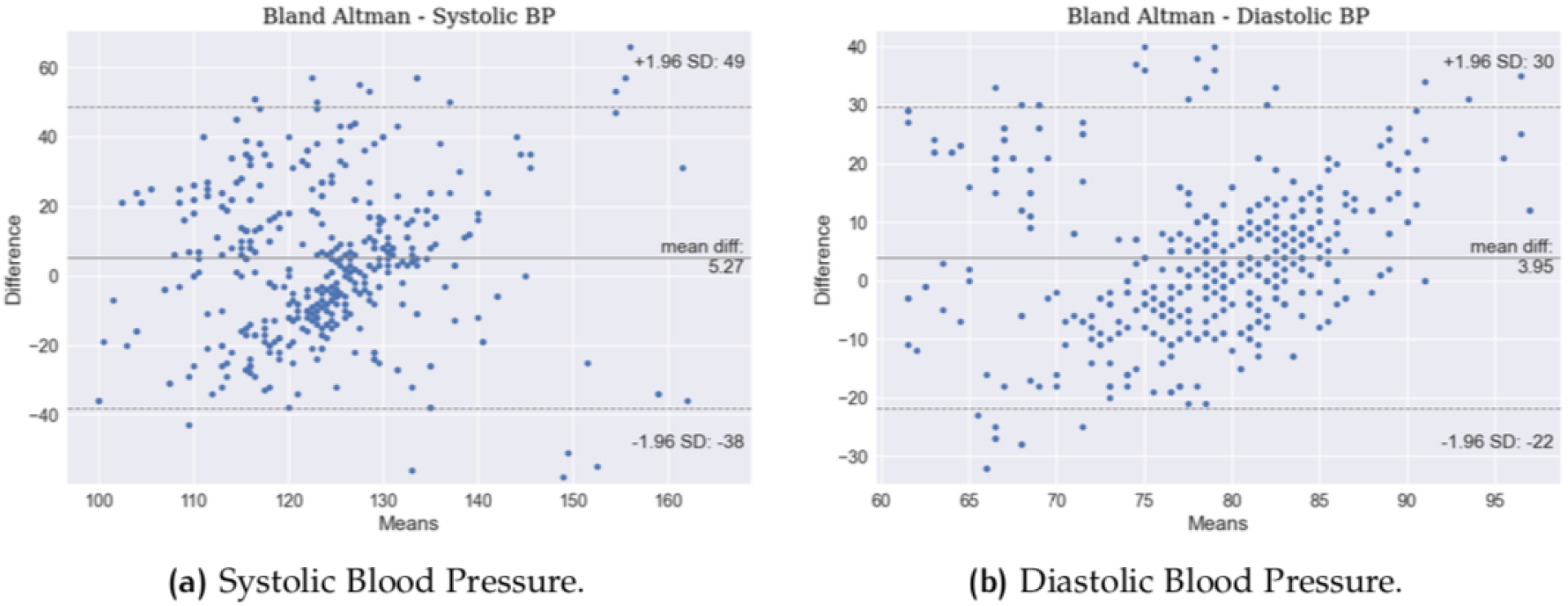
Bland Altman Plot for: a) Systolic and b) Diastolic

This study proposes a deep dive into the presented results, so, Fig.8 evaluates the impact of age on the errors. Similarly, Fig.9 shows the influence of height and weight.

**Figure 8:**
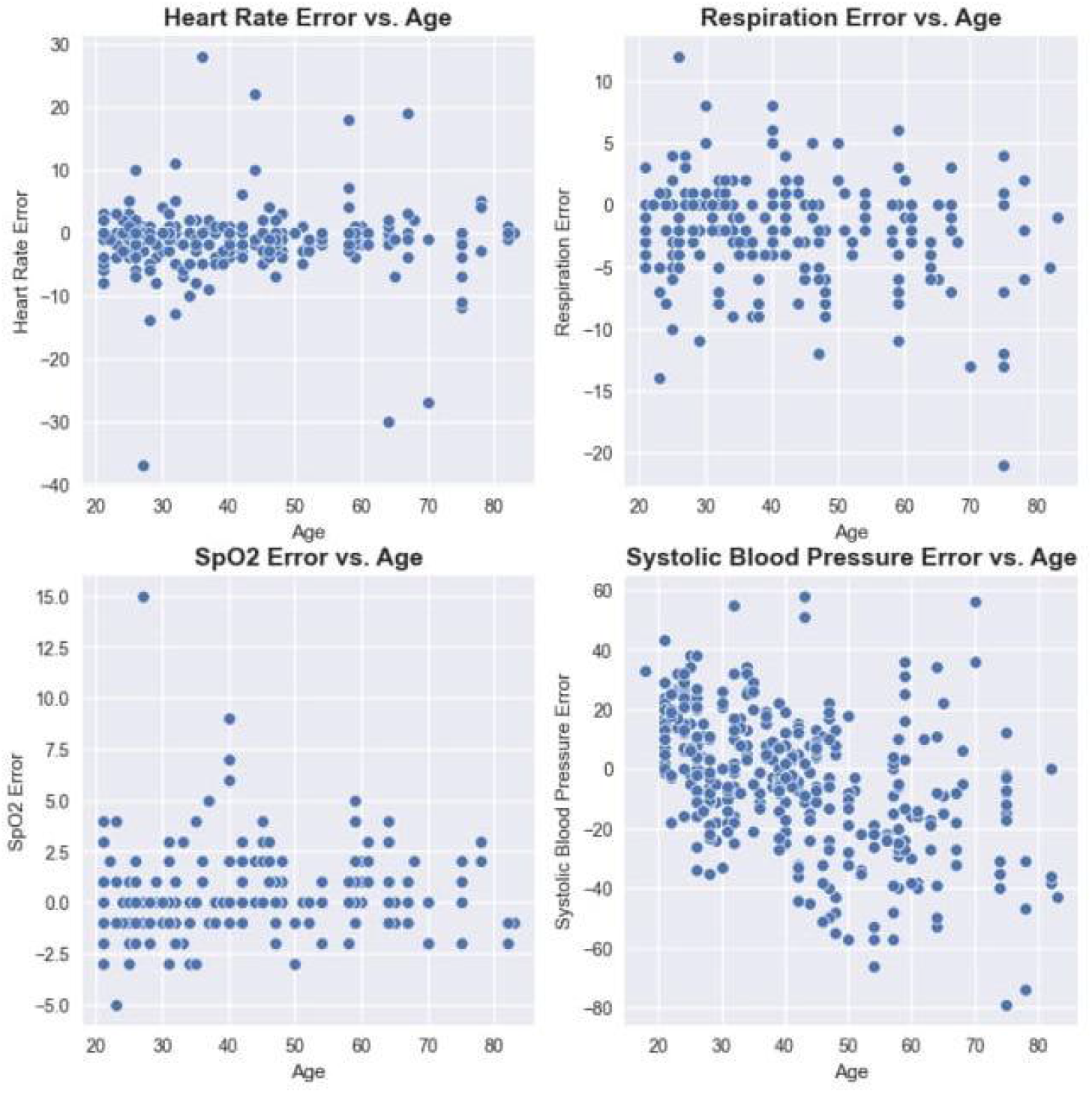
Impact of age in vitals errors for Heart Rate, Respiration, SpO2 and Systolic Blood Pressure.

**Figure 9:**
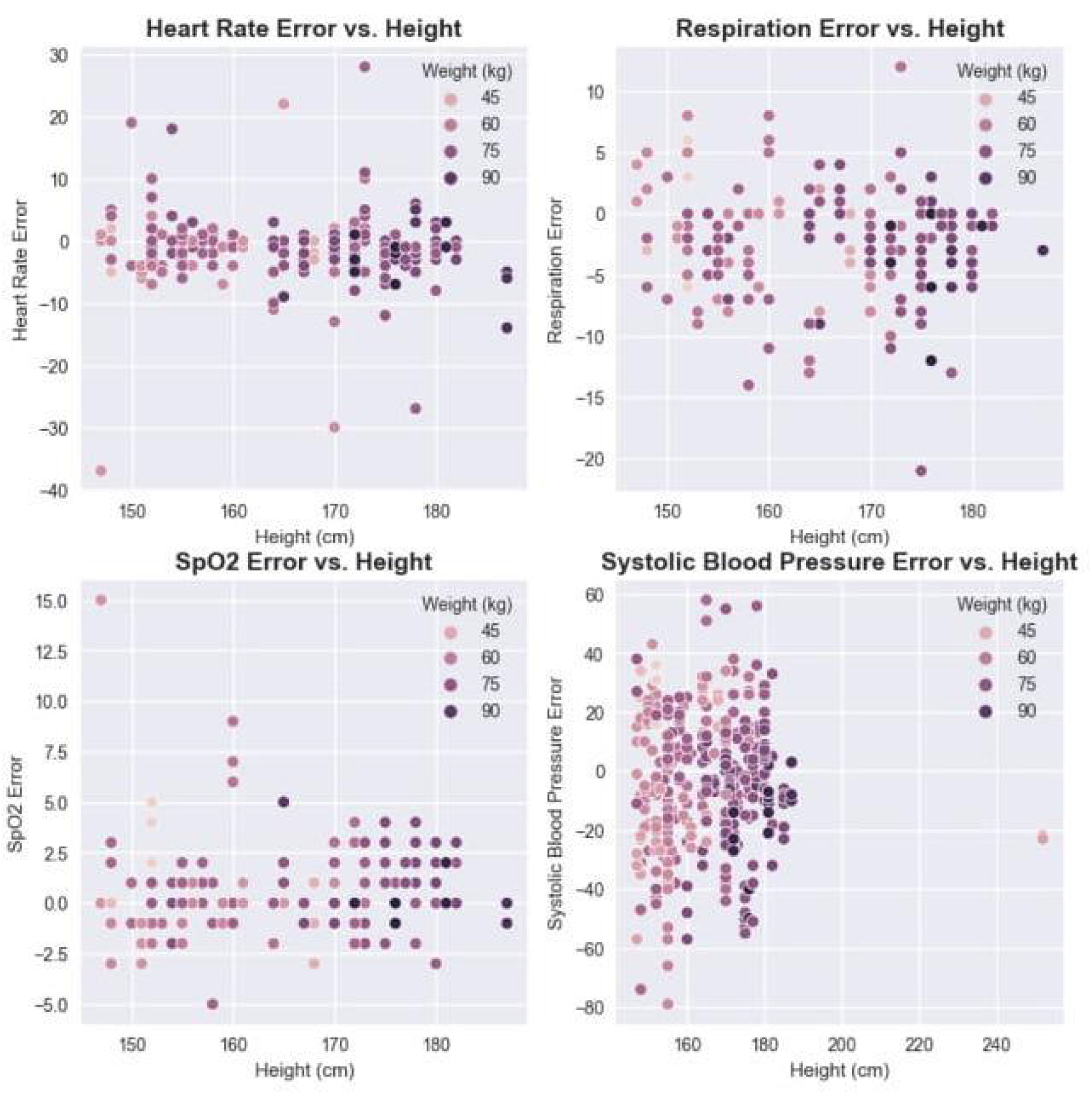
Impact of height and weight in vitals errors for Heart Rate, Respiration, SpO2 and Systolic Blood Pressure.

Furthermore, it is well known that there are going to be outliers, which might drive the statistical results away. Through box plots, this study aims to show the quartile ranges as well as the outliers for each one of the vitals, described in Fig.10.

**Figure 10:**
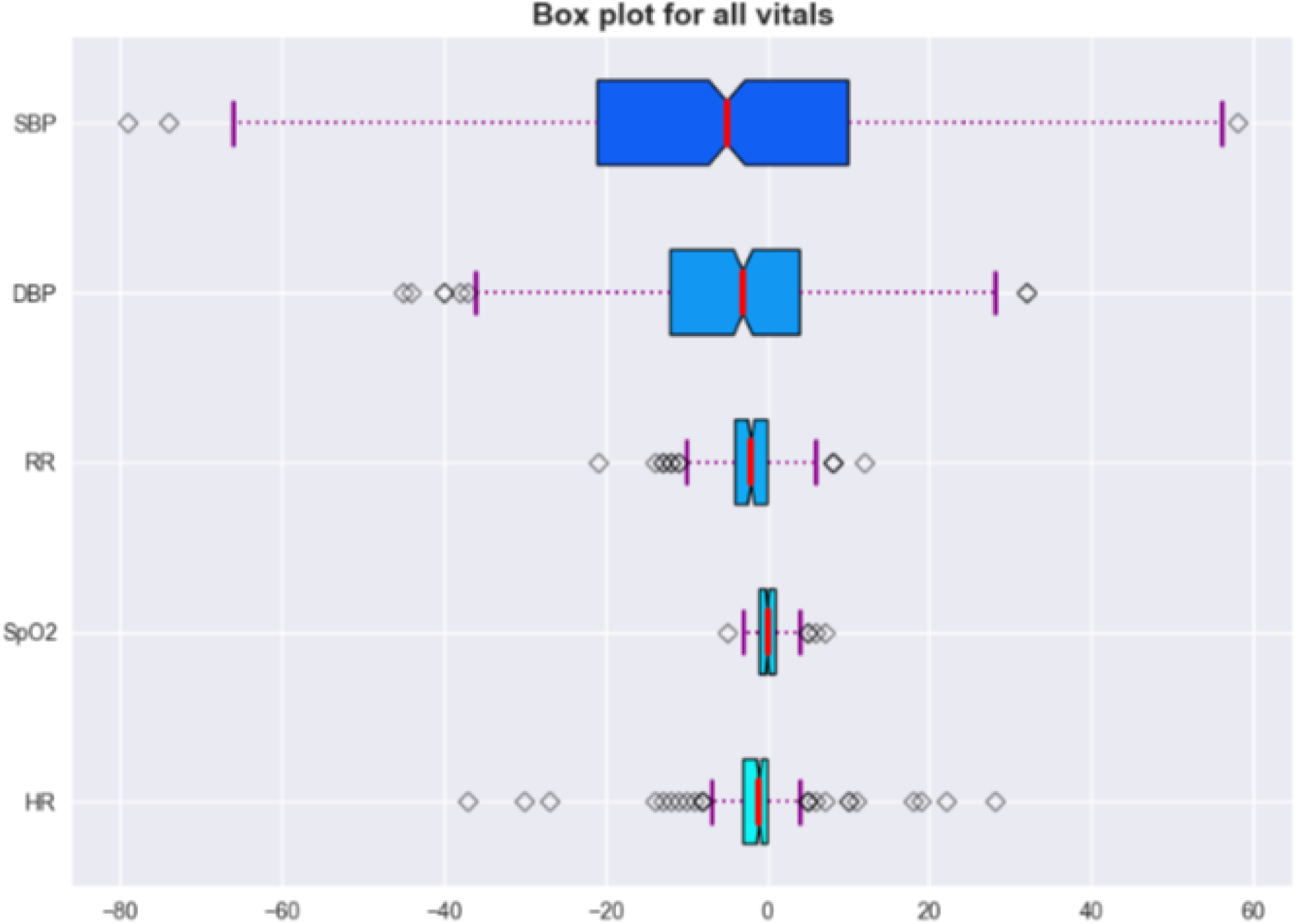
Box plot for all vitals.

## DISCUSSION

The present study aims to evaluate visual technology against medical devices. In this study, technology is subject to frequent software upgrades to continuously improve its ability, therefore the performance of the tool in studies conducted recently is likely to outperform older studies. In this study, data from 463 subjects have been collected, however, not all the vitals were collected for each one of the patients. Blood Pressure is the only one always present. The sample size of the others was decreased to around 240 patients [10-11].

Overall, the results show good agreement between camera based visual monitoring technology and the reference instruments. Heart Rate, Respiration, and Oxygen Saturation reached the hypothesis criteria of +/-3 units in mean error for each parameter. Systolic and Diastolic Blood Pressure also did meet the requirements of +/-10 units in mean error, however, this should be noted with caution given the high standard deviation of error amongst the results [12].

With regards to the other metrics, it is possible to see that using MAE most of the algorithms were able to keep similar results, apart from Systolic Blood Pressure, which highly increased in absolute difference. When applying a quadratic function, such as RMSE, it is possible to highlight errors that would be smoothed by the mean value. Having said that, RMSE from heart rate and SpO2 show acceptable results in the average and within each group, however, in respiration rate and further in SBP and DBP it is possible to see the impact of large errors in a quadratic metric [13].

One of the most robust algorithms is indeed the heart rate estimation, which was able to keep +/-3 criteria in the average as well as within low and normal ranges. Naturally, the extreme cases of either high or low heart rate imply more difficulty to estimate, especially higher values that can be associated with motion artifacts due to the stress that the patients might face, for obvious reasons of being at a hospital or just for participating in the research study.

Oxygen Saturation (SpO2) shows acceptable agreement between 90% and 100%, with a 0.3% of mean error. In this study, there were limited samples with SpO2 below 90%, since these cases usually only occur for individuals in either a critical situation or high altitude setting and future work will look to include more participants with hypoxia to validate the agreement at such low levels. Nevertheless, it is worth mentioning the current scope of the technology which will more likely be used to prevent such conditions rather than to be used during it [14].

Respiration rate, on other hand, has shown the best performance in the normal respiration range between 10 and 18 breaths per minute, with a mean error of -0.48. The highest bias can be observed in very low levels, less than 10 breaths per minute.

Moreover, Blood pressure, in terms of Systolic and Diastolic is the most challenging problem to solve, in this study, the technology used has shown good results, considering the results and the number of samples within each group, however struggling with extreme values, mostly in SBP higher than 150 mmHg and DBP higher than 90 mmHg [15].

Considering the histograms from Fig.4 and Fig.5 it is possible to see the tendency of narrowing the plot close to error around 0, which shows that the most frequent results have lower levels of error. However, outliers might impact the statistical results. The heart rate histogram Fig.4c shows one perfect example with the classic normal (bell pattern) distribution [16].

Bland Altman plots provide a more robust view than linear scatter plots since it is possible to see the results in terms of mean value vs. difference. Fig.6c shows that the mean difference for HR is 1.19 bpm, with a higher standard deviation of +12 -10, while RR shows a lower SD with a higher mean difference, but with more sparse errors. SpO2 as should be, displays fewer data points because more than one reading had the same behavior (estimated and reference), which leads to a lower mean difference since the results are grouped. Bland Altman plots for Blood Pressure show more consistently a pattern of the wrong prediction in extreme cases with SBP lower than 120 mmHg and higher than 140 mmHg, similarly, DBP lower than 75 mmHg and higher than 85 mmHg. These results show the expected behavior for this version of Blood Pressure, which can generalize very well within a normal range of BP and struggle with extreme cases, both low and very high values [17].

To investigate whether age has a strong role in the error or not, this study has shown in Fig.8 that no correlation between age and error can be observed in any one of the vitals. This is meaningful once no age bias is present and can be used by patients in any part of their lives. Moreover, neither height nor weight seems to influence the results as per Fig.9 which shows that the relationship between body weight and height (Body Mass Index-BMI) has no visible effect in the presented results. Nevertheless, the results show that it does not suffer from bias related to specific groups of demographic characteristics, such as age, gender, height, or weight [18].

A Box plot shows the errors in terms of their distribution into quartiles as well as the outliers. As can be seen in Fig.10 HR, RR and SpO2 show low interquartile ranges, while this range increases in DBP and further in SBP. These results meet the initial observations that blood pressure has a higher standard deviation in comparison with the other vitals. In addition, it is also possible to see some outliers in Heart Rate estimation which can drive the statistical results and may be related to wrong ground truth measurement. Yet, heart rate was able to achieve acceptable results even though these outliers might be present [19].

## CONCLUSION

In this study, 463 readings were used to analyze to investigate the accuracy of visual technology. The study was conducted in India at multiple facilities as well as in the United Kingdom with Leeds Teaching Hospital Trust with patients with a variety of demographic characteristics.

The study has shown that the technology meets acceptable agreement levels for mean error for HR, RR, SpO2, and BP, however, large deviations were apparent across several BP readings. This shows that the use of camera based monitoring solutions are acceptable for users who want to understand their general health and wellness.

## Data Availability

Researchers interested in access to the data may contact Nikhil Sehgal at, also see https://vastmindz.com/contact/. The authors will assist with any negotiable data use agreements and gain access to the data for any replication efforts following publication.

## ETHICS APPROVAL

The study was carried out in multiple facilities:

- Ayurvedic and Unani Tibbia college, New Delhi, India, 100 subjects Ethics Committee approved.
- PSRI Hospital and Research Institute, New Delhi, India, 100 subjects Ethics Committee approved.
- Fortis Hospital, Noida, India, 275 subjects Ethics Committee approved.
- Leeds Teaching Hospital Trust, United Kingdom, 27 subjects, Ethics Committee approval from UoL.

All subjects provided written informed consent prior to participation in the study.

